# Do Researchers Consider the Inter-relationship Between Time to Assessment and Admission Severity in Acute Stroke?

**DOI:** 10.1101/2024.11.28.24318128

**Authors:** Zewen Lu, Matthew Gittins, Amit K Kishore, Craig J Smith, Andy Vail

## Abstract

**Background:** Stroke severity, often quantified by NIHSS at admission, evolves and may influence patients’ time to hospital arrival. The inclusion of timing in stroke severity assessment remains unclear in stroke research. Oversight of time to assessment can affect prognostic model interpretation, acute clinical decision-making, and the design of future clinical trials by considering admission severity in the context of time since symptom onset.

**Aims:** This study aimed to assess whether and, if so, how stroke researchers account for time from symptom onset to admission severity assessment in their analyses. We sought to compare this with approaches used by perinatal researchers, considering a similar statistical relationship between gestational age and birth weight.

**Summary of review:** Two reviewers systematically reviewed papers in leading specialty journals published in 2019 using NIHSS at admission and birth weight respectively as an explanatory factor in the statistical analysis. We targeted a minimum of 50 articles from each field to ensure 90% to identify approaches used in 5% or more of studies.

A total of 111 studies were included. Perinatal researchers considered the temporal variable gestational age more often than time to assessment in stroke studies (89% vs. 7%, chi-squared p<0.001). It was consequently included more often (56% vs. 5%, chi-squared p<0.001). Four methods, including stratification, distribution, regression and combined approaches, were found. Time to assessment was only included as a continuous (n=2) or categorical (n=1) factor alongside admission NIHSS in three studies. Methods for covariate selection, essential for the interpretation of statistical models, were rarely specified.

**Conclusions:** Few researchers explore the inter-relationship between baseline severity and time to assessment in stroke prognostication, in sharp contrast to consideration of timing in perinatal studies. Future research will investigate whether time to assessment or serial NIHSS in the hyper-acute phase could benefit both clinical practice and stroke research.

## 1 Introduction

Stroke may progress rapidly in the first few hours following ictus, leading to severe complications if left untreated, underscoring the mantra “time is brain” [^1^]. The severity of stroke on admission to the hospital is typically measured by the National Institutes of Health Stroke Scale (NIHSS), a 15-item examination capturing various neurological deficits post-stroke [^2 3 4^]. Admission severity is a crucial prognostic marker for outcomes such as stroke-associated pneumonia, discharge destination, and functional recovery [^5 6 7 8 9^]. Adjusting for admission stroke severity is widely recognised for improving model fit and predictive power. [^10^].

Stroke symptom severity can evolve rapidly, particularly within the first few hours, affecting the recorded severity at the time of hospital admission [^11^]. Additionally, while all stroke survivors are advised to seek immediate medical attention, actual times to admission vary widely [^12^]. Patients with more severe symptoms may arrive earlier, suggesting that stroke severity influences the time to assessment.

Given the dynamic nature of stroke progression, incorporating longitudinal designs with repeated measures could enhance prognostication [^13 14^]. Including a dynamic, time-varying covariate as a fixed baseline measure in statistical models can also lead to biased estimates of patient survival [^15^]. However, in clinical practice, stroke severity is often recorded only once at admission [^16^]. This still creates a complexity: controlling for the timing of assessment if the timing itself is influenced by the evolving symptom severity.

We, therefore, systematically reviewed recent high-profile journal publications to determine if, and how, time to assessment is incorporated into studies using admission severity as a prognostic factor. To provide a comparative perspective, we looked at perinatal research, which deals with similar statistical challenges. Perinatal studies frequently examine the relationship between gestational age at delivery and birthweight, akin to the relationship between time to assessment and stroke severity. This field has established robust methods for adjusting for the timing of measurements, such as stratifying outcomes by birthweight and gestational age, including both variables in analyses, and defining birthweight centiles by age [^17 18 19 20 21 22 23 24 25 26^].

Time (gestational age) adjustment in perinatal research allows for a more accurate assessment of a newborn’s growth and health status. Given the analogous nature of the data structures— both involving an evolving condition (fetal growth or stroke severity) measured at a time-dependent point (gestational age or time to hospital admission)—it is plausible that similar approaches could be beneficial in stroke research by adequately accounting for the timing of admission severity assessments.

## 2 Methods

### 2.1 Study design and aim

We undertook a systematic (protocolised) review of methodology, specifying strict eligibility criteria. Our interest was focused on the statistical approach, if any, taken to address the inter-relationship between the time to assessment (onset to admission time or gestational age) and the realisation of the evolving prognostic factor at that time-point (admission NIHSS or birthweight). The clinical question and results were not of interest so therefore did not attempt meta-analysis or quality assessment of the included studies.

We followed the PRISMA guidelines to ensure the quality and transparency of reporting of a systematic review [^27^]. The protocol of this review was prespecified but not registered. It was not published but can be provided if requested.

### 2.2 Sample size calculation

For comparison of the two research fields, we suspected that the inter-relationship would be considered in at least 80% of the perinatal papers and no more than 20% of stroke papers.

Such a difference could be detected with 90% statistical power with only 12 papers from each field. We chose to study 50 from each in order to achieve over 90% power to find a method used in only 5% of papers.

### 2.3 Search and screening strategy

Based on the sample size calculation, we chose to review leading specialty journals for a single year. We chose 2019 as the most recent year in which the clinical research would not have been affected by the COVID-19 pandemic. We chose journals based on the ranking of journal citation reports (JCR) 2021 [^28^] and the number of published articles identified by a keyword search.

The initial searching and eligibility criteria were piloted by two independent reviewers (ZWL and AV). The initial screening was based on searching keywords ‘NIHSS OR Severity’ in stroke journals and ‘Weight OR Birthweight’ in perinatal journals. We further screened titles and abstracts to exclude non-clinical and secondary studies.

### 2.4 Eligibility criteria

We included studies incorporating an analysis of the association between admission severity (or birthweight) and subsequent clinical outcomes, regardless of study design. We excluded studies with fewer than 40 participants as muti-variable regression may not have been appropriate [^29^]. We also excluded studies that only analysed serial observations of NIHSS (or fetal weight) as such analyses intrinsically recognise the dynamic nature of the prognostic factor using data not available in our motivating context.

### 2.5 Data extraction and analysis

We extracted data using a bespoke form in Microsoft Excel. We recorded information on publication characteristics, study design and role of admission NIHSS (or birthweight) in the analysis. For method comparison, we also recorded the statistical analysis methods, the outcome of interest, and any covariate selection procedure.

We first summarised the characteristics of the included studies. We compared whether ‘time to assessment’ (or gestational age) was considered in each study using a simple chi-squared test and catalogued how, noting when multiple eligible approaches were presented. Finally, we tabulated methods of covariate selection as these might have affected the inclusion of the temporal variable ‘time to assessment’ or ‘gestational age’.

## 3 Results

### 3.1 Systematic search and study selection

The top two journals in the perinatal field had 132 articles containing our keywords (51 Neonatology; and 81 Journal of Perinatology). We selected the first and sixth journals in the stroke field (32 Stroke; 61 Journal of Stroke and Cerebrovascular Disease) as the intervening journals had lower numbers of such articles. After eligibility screening (Figure 1) we included a total of 111 papers, 54 (49%) studies in the perinatal group and 57 (51%) in the stroke group.

**Figure 1.**
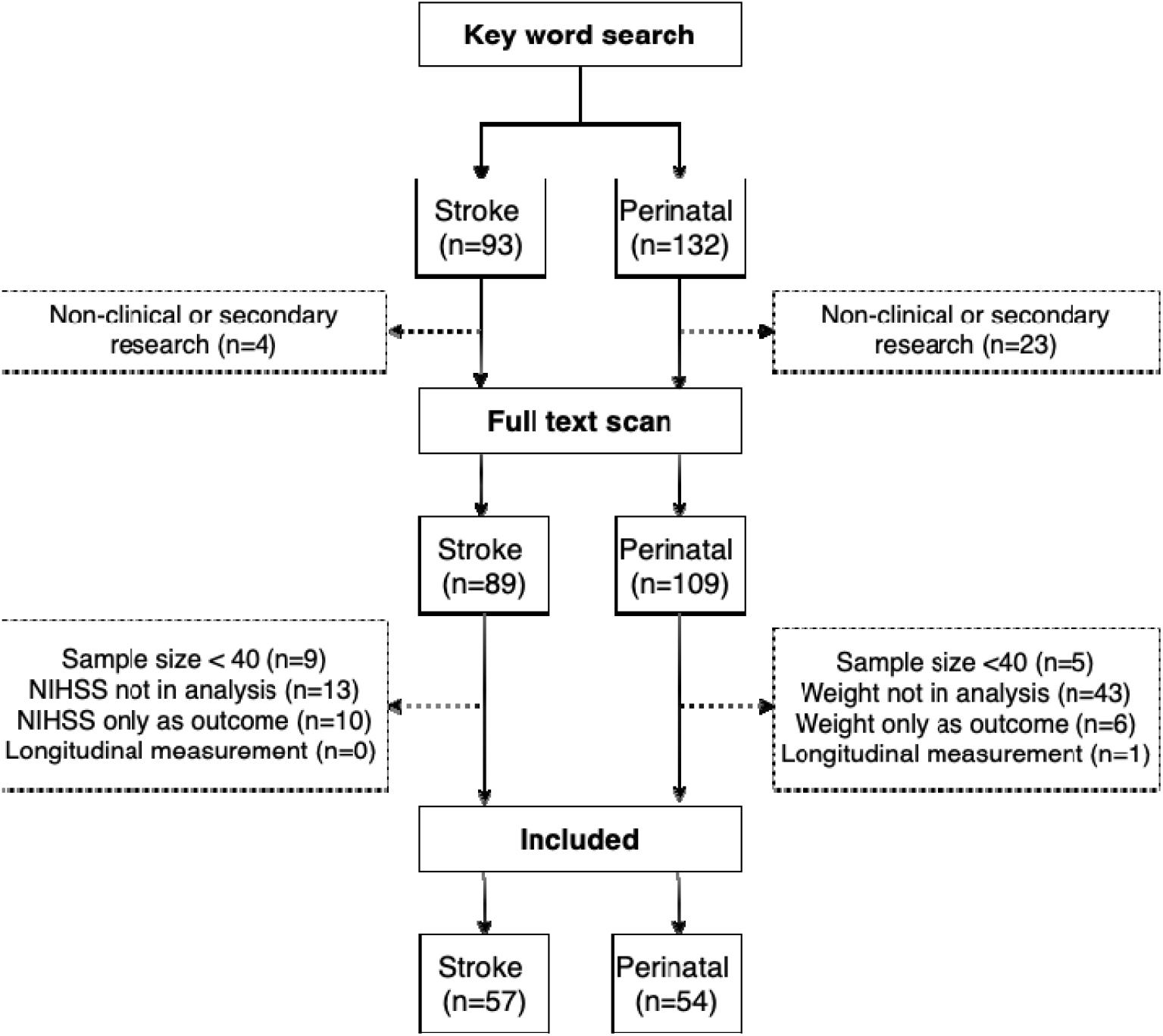
Flow chart of the study selection

**Figure 2.**
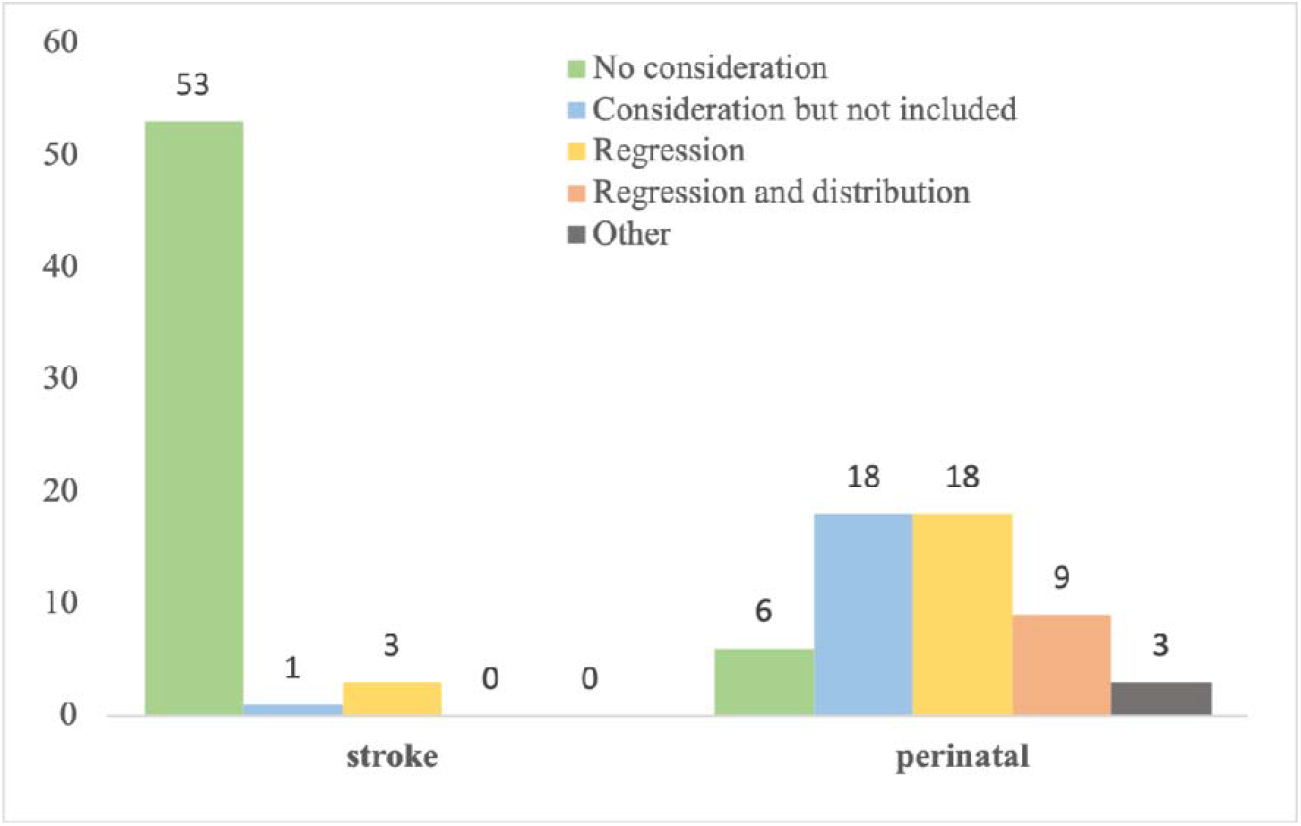
Frequency of methodology to incorporate the dynamic inter-relationship between stroke and perinatal groups

### 3.2 Study characteristics

The characteristics of the included studies are summarised in Table 1. Both fields were dominated by North American studies. A minority clearly identified a statistician within the authorship, but this was more common in the stroke field. Perinatal studies focused more on risk factor assessment but the majority of papers in each field were accounting for the dynamic marker as a confounder. Studies in stroke were less likely to restrict their eligibility criteria by either factor.

**Table 1.**
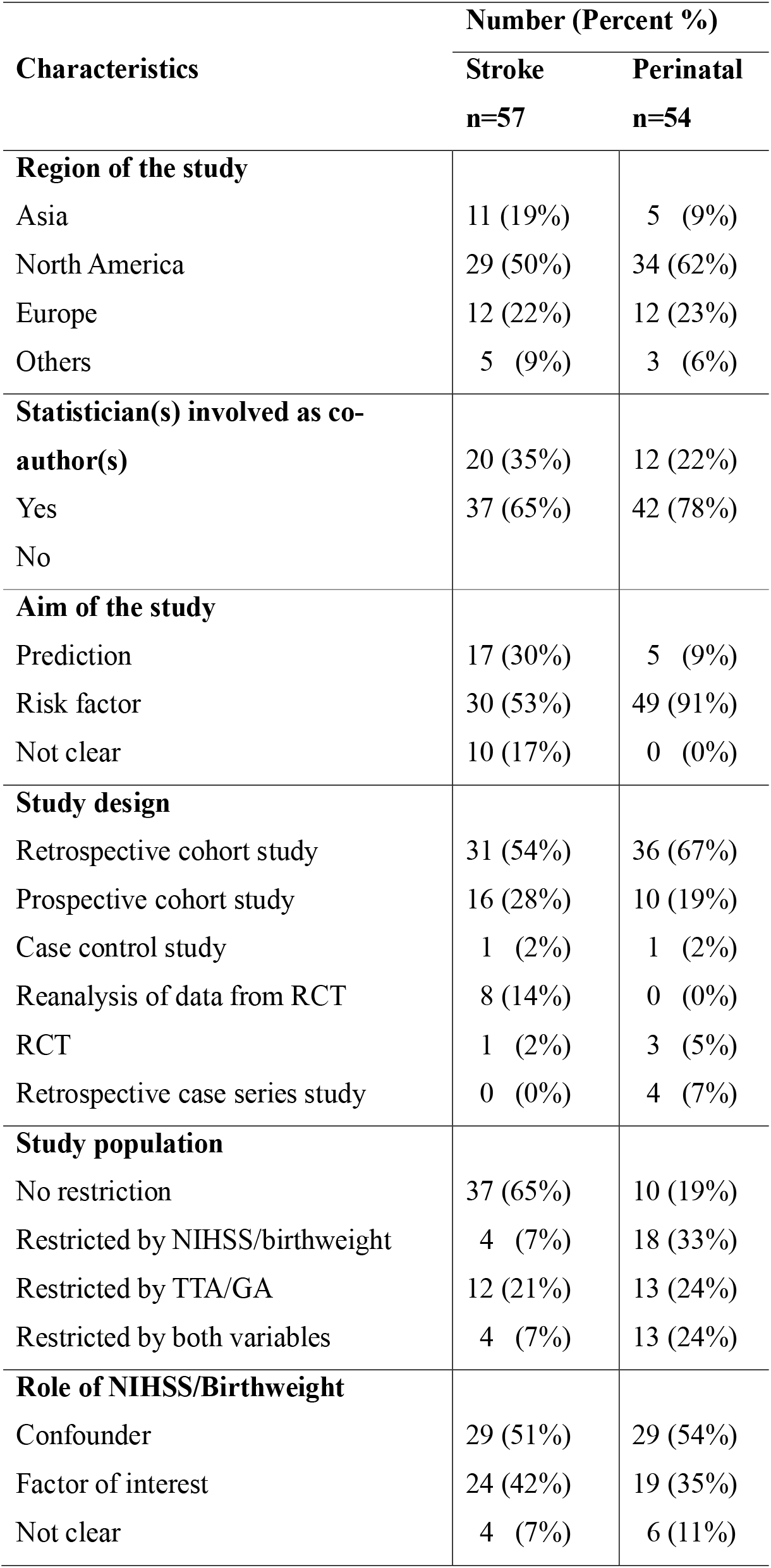
Summary of characteristics of included studies.

### 3.3 Methodology comparison

#### 3.3.1 Overall statistical analysis

Both fields reported mostly binary outcome variables and regression models (Table 2). Most studies did not report the methods used to select covariates. For those that did, there was a wide variety of different approaches. Interestingly, the relatively modern statistical approach of propensity score models [^30 31^] was identified in stroke (three papers) but not perinatal research. Those using this approach all chose to include admission NIHSS as a matching factor. They used matching, inverse probability of treatment weighting (IPTW) or both as the method for estimate adjustment. Only one study included both admission NIHSS and time to admission as matching covariates to estimate the propensity score.

**Table 2.** Summary of overall statistical analysis for the included studies.

| Statistical analysis | Number (Percent %) |  |
| --- | --- | --- |
|  | Stroke<br>n=57 | Perinatal<br>n=54 |
| **Outcomes** |  |  |
| Multiple | 8 (14%) | 4 (7%) |
| Continuous | 5 (9%) | 14 (26%) |
| Binary | 43 (75%) | 34 (63%) |
| Ordinal | 1 (2%) | 0 (0%) |
| Time to event | 0 (0%) | 2 (4%) |
| **Statistical analysis** |  |  |
| Matched in design | 2 (4%) | 0 (0%) |
| Included in regression | 52 (90%) | 44 (81%) |
| Both design and regression | 1 (2%) | 0 (0%) |
| Statistical test | 1 (2%) | 8 (15%) |
| Summary statistics | 1 (2%) | 2 (4%) |
| **Covariate Selection** |  |  |
| Baseline characteristics | 1 (2%) | 1 (2%) |
| Literature review and clinical relevance | 3 (5%) | 2 (4%) |
| Stepwise forward selection | 1 (2%) | 1 (2%) |
| Stepwise backward selection | 5 (9%) | 2 (4%) |
| Bi-directional stepwise selection | 2 (4%) | 1 (2%) |
| Univariable analysis | 13 (23%) | 4 (7%) |
| Multistage selection | 5 (8%) | 5 (9%) |
| Unspecified | 27 (47%) | 38 (70%) |

#### 3.3.2 Consideration of dynamic inter-relationship

As anticipated, most papers in the perinatal field (89%) and very few (7%) in stroke indicated some consideration of the inter-relationship by incorporation of the temporal factor alongside the dynamic marker (chi-squared = 71.39, p<0.0001, Table 3). Consequently, the temporal variable was also included more often (56% vs. 5%, chi-squared=31.2, p<0.0001, Table 3).

**Table 3.**
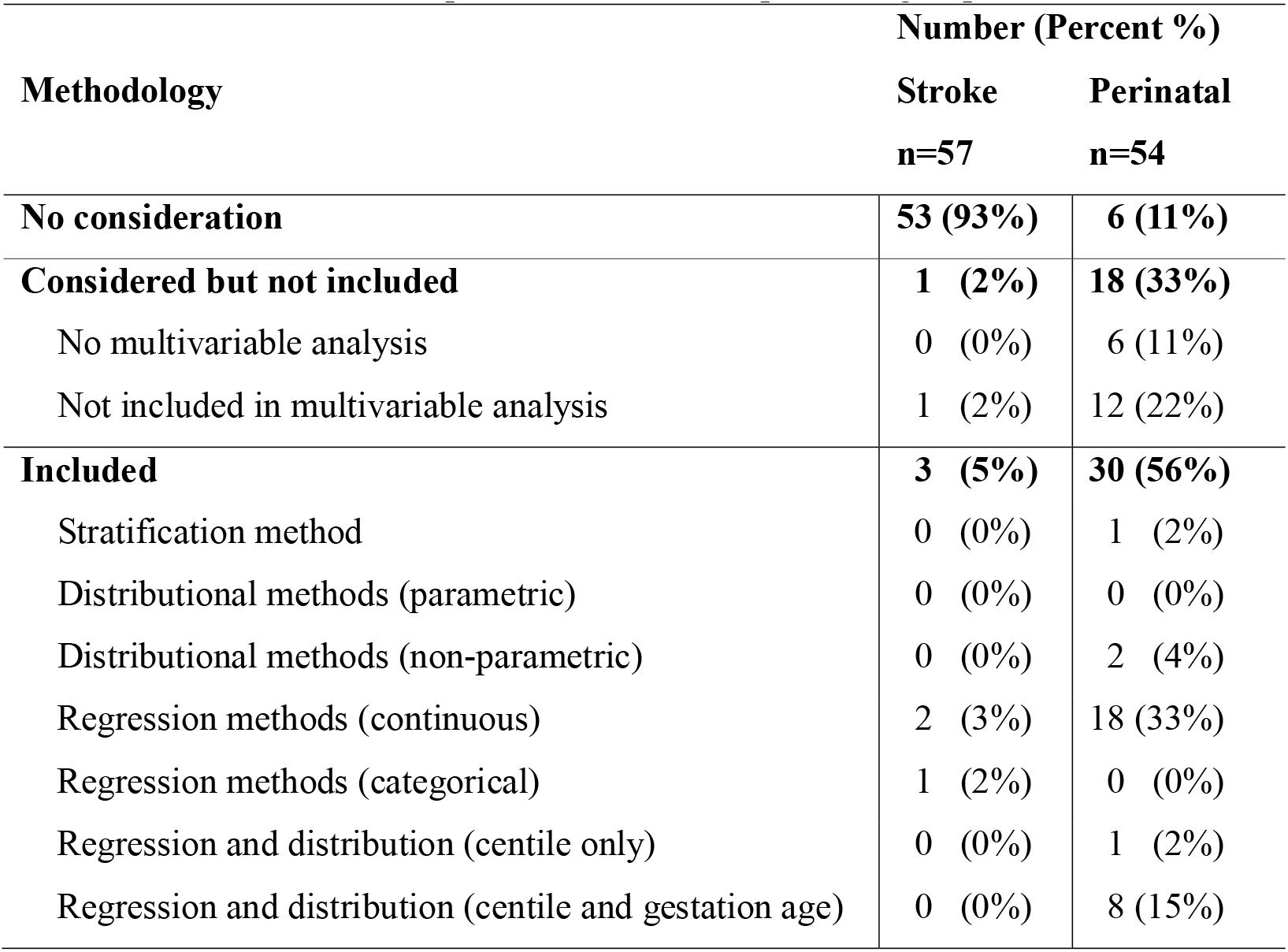
Summary and comparison of methodology to incorporate the dynamic inter-relationship between stroke and perinatal groups.

| Methodology | Number (Percent %) |  |
| --- | --- | --- |
|  | Stroke<br>n=57 | Perinatal<br>n=54 |
| **No consideration** | **53 (93%)** | **6 (11%)** |
| **Considered but not included** | **1 (2%)** | **18 (33%)** |
| No multivariable analysis | 0 (0%) | 6 (11%) |
| Not included in multivariable analysis | 1 (2%) | 12 (22%) |
| **Included** | **3 (5%)** | **30 (56%)** |
| Stratification method | 0 (0%) | 1 (2%) |
| Distributional methods (parametric) | 0 (0%) | 0 (0%) |
| Distributional methods (non-parametric) | 0 (0%) | 2 (4%) |
| Regression methods (continuous) | 2 (3%) | 18 (33%) |
| Regression methods (categorical) | 1 (2%) | 0 (0%) |
| Regression and distribution (centile only) | 0 (0%) | 1 (2%) |
| Regression and distribution (centile and gestation age) | 0 (0%) | 8 (15%) |

Of only four (7%) papers in the field of stroke that considered time to assessment, one did not retain it, three included it in the statistical analysis using regression methods, two as continuous (assumed linearity) and one as categorical (dichotomised at 360 mins).

In contrast, only six (11%) studies in the perinatal field did not consider gestational age including one that specified the data were unavailable. Those that did used a variety of methods including a simple regression method that treated gestational age as a continuous explanatory factor, centile methods that defined a binary variable as ‘small for gestation age’ (SGA) or ‘large for gestational age’ (LGA) in regression, a χ^2^ test or Fisher’s exact test or just summary statistics of the outcome between two groups by the dichotomised birthweight & GA, SGA and LGA.

## 4 Discussion

This review has highlighted that researchers in the field of stroke rarely recognise that symptom severity in the hours following a stroke is a dynamic process rather than a fixed baseline characteristic. Despite the critical importance of time to diagnosis and treatment in acute stroke management, the relationship between baseline severity and time to assessment was not widely considered in our review of stroke research [^32 33^].

The inclusion of time to measurement for diagnostic tools or treatments may depend on its association with patient outcomes [^34 35^]. We may assume that early diagnoses and treatments lead to a better prognosis. One conventional thinking is that the delay caused by organisational or socioeconomic defects has an adverse impact on patient outcomes. For instance, logistical challenges faced by patients living remotely or alone can delay hospital admission, even amidst acute symptom onset [^36 37^]. However, the initial assessment of stroke severity cannot diagnose a stroke. Therefore, researchers may not relate time to assessment with patient outcomes. This may explain the lack of attention to incorporating time to assessment or its inter-relationship with admission severity in stroke research.

Besides, the magnitude of association between admission severity and time to assessment is also mild, with a negative linear correlation of -0.2 during the initial 4 hours and tapering to - 0.06 thereafter [^11^]. Contrastingly, in the example of perinatal research, the positive correlation between birthweight and gestational age is stronger and more consistent. Few researchers considering birthweight attempted to do so in the absence of information on gestational age. Extensive research in perinatology has yielded birthweight centile charts, facilitating classifications such as small for gestational age and large for gestational age [^38 39^]. These charts further account for variables like maternal anthropometrics, parity, and ethnic backgrounds. Furthermore, birth outcomes pivoting around these classifications, both population-centric and custom-tailored, have been comparably studied [^40 41^]. We are unaware of any corresponding considerations in stroke, such as “mild for 1-hour admission”. This might help better define the dynamic symptom severity, leading to an optimal interpretation of stroke prognostication. However, there is also a lack of evidence in the design of clinical trials or clinical practice to distinguish patients with the same admission NIHSS scores but different assessment/admission times. Trials select eligible patients based solely on admission severity without considering the time of measurements or any changes in symptom severity before randomisation [^42^].

The procedure of covariate selection may also impact the inclusion of time to measurement in prognostic models. Gestational age is widely considered a prognostic marker in its own right, whereas time to assessment is less recognised as such and rarely included in the table of baseline characteristics. Researchers selecting on the basis of literature review are therefore less likely to include time to assessment as a covariate. Similarly, if researchers select covariates based on their uni-variable statistical significance with outcome, they may be less likely to include time to assessment due to the weaker relationship. It is less clear what the effect of stepwise selection strategies may be. The weaker association between time to assessment and subsequent outcome may lead to exclusion. On the other hand, gestational age may also be excluded due to high correlation and multicollinearity with birthweight. The choice of the threshold of p-value in the stepwise selection and univariable analysis plays an important role in the temporal variable to be included in the multivariable model. Despite its central role, most studies in this review did not specify the method of covariate selection.

We restricted our review to only two leading journals in each group for a single year. We make no claim that this review is comprehensive nor do we seek to estimate the exact proportion or difference in the proportion of researchers considering the temporal variables between the stroke and perinatal fields. However, we deem it unlikely that more sophisticated methods of analysis would be employed in journals of lower citation rank. Equally, this is not a comprehensive review of the methods available to incorporate the inter-relationship of either gestational age and birthweight or time to assessment and admission severity. Although the sample size calculation ensures a high chance of finding methods applied in these fields, other methods may be applied elsewhere or developed in the statistical literature but yet to be applied.

## 5 Conclusion

The timing of admission severity is rarely considered in stroke, in sharp contrast to the timing of birthweight in the perinatal field. Given the dynamic state of symptom severity, clinicians may consider admission severity in the context of time since symptom onset and the value of repeat NIHSS assessment before the end of any time window for acute intervention. For epidemiologists, there may be reasonable statistical justification for this lack of consideration related to the inconsistency of the evolution of symptoms in the hours following stroke and the strength and shape of the relationship between this evolution and clinical outcomes. However, with a better understanding of these factors, there is potential to improve both clinical care and stroke research.

## Data Availability

All data produced in the present study are available upon reasonable request to the authors

## Notes

### Competing Interest Statement

The authors have declared no competing interest.

### Funding Statement

This study did not receive any funding

